# Substance Use Onset in High-Risk 9-13 Year-Olds in the ABCD Study

**DOI:** 10.1101/2021.08.10.21261808

**Authors:** Natasha E. Wade, Susan F. Tapert, Krista M. Lisdahl, Marilyn A. Huestis, Frank Haist

**Affiliations:** Department of Psychiatry, University of California, San Diego; Department of Psychology, University of Wisconsin-Milwaukee; Institute of Emerging Health Professions, Thomas Jefferson University

**Author notes:** Correspondence and reprint requests to: Natasha E. Wade, Ph.D., Assistant Professor, Department of Psychiatry, University of California, San Diego, 9500 Gilman Drive, MC 0405, La Jolla, CA 92093.

**Keywords:** children, adolescents, hair toxicology, substance use, substance use onset, self-report, hair samples

## Abstract

**Aim:** A key aim of the Adolescent Brain Cognitive Development□ (ABCD) Study is to document substance use onset, patterns, and sequelae across adolescent development. However, substance use misreporting can obscure accurate drug use characterization. Hair toxicology provides objective historical substance use data but is rarely used in studies of youth. Here, we compare objective hair toxicology results with self-reported substance use in high-risk youth.

**Methods:** A literature-based substance use risk algorithm prioritized 696 ABCD Study^®^ participants for hair sample analysis at baseline, and 1 and 2-year follow-ups (spanning ages 9-13). Chi-square and t-tests assessed differences between participants’ demographics, positive and negative hair tests, risk-for-use algorithm scores, and self-reported substance use.

**Results:** Hair testing confirmed that 17% of at-risk 9-13 year-olds had evidence of past 3-month use of one (n=97), two (n=14), three (n=2), or four (n=2) drug classes. After considering prescribed medication use, 10.3% had a positive test incongruent with self-report. No participant with a positive result self-reported recreational substance use that was consistent with their toxicology results. They also reported less sipping of alcohol (*p* < 0.001) and scored higher on the risk-for-use algorithm (*p* < 0.001) than those with negative toxicology results.

**Conclusions:** 10% of at-risk 9-13 year-olds tested positive for at least one *unreported* substance, suggesting underreporting in this age range when participating in a research study. As hair testing prioritized youth with risk characteristics, the overall extent of underreporting will be calculated in future studies. Nonetheless, hair toxicology was key to characterizing substance use in high-risk youth.

---

The Adolescent Brain Cognitive Development□ (ABCD) Study is a landmark project of healthy development following primarily substance-naïve youth (9-10 years-old) prospectively for 10 years with the expectation that a proportion will initiate substance use [1]. The neurocognitive, psychosocial, psychiatric, and neurobiological predictors and sequelae of substance use are evaluated. The ABCD Study^®^ employs the Timeline Follow-Back [2] gold standard for reporting substance use, a self-report semi-structured interview where participants are guided to recall their past year’s substance use patterns including dose, estimated potency, and routes of administration [1].

Retrospective self-report has significant limitations. Lack of accuracy, regardless of participant age, may be due to intentional or unintentional misreporting, due to perceived desirability, concerns regarding privacy [3, 4], forgetting, and lack of knowledge of substances taken [3]. Some factors influencing reporting accuracy include demographics [5], mental health [6], age of first use [6], frequency of use [5], and guidance regarding standard units [7]. Age may also influence substance use disclosures, though this was not investigated and the accuracy of adolescents’ self-reported substance use is not well understood. Many prior youth studies were conducted in treatment or juvenile detention facilities or as part of a community-based substance use study, settings carrying differential motivation to misrepresent their use. One study of a generally healthy population of male adolescents whose fathers had alcohol dependence revealed a 13% discordance rate between adolescent self-report and urine drug screen results [4]. Recently, advanced analyses utilizing data on misreporting estimated that self-reported cannabis use in Washington State adolescents is underreported, with 7% who took cannabis in the past month denying use [8]. A larger characterization of self-report accuracy in relation to objective substance use measures is important, particularly in adolescents.

Though the Timeline Followback was validated with test-retest assessment and toxicology in adults [9-11], a new gold standard of combining self-report with objective measurement was proposed [11]. Accordingly, the ABCD Study was designed to measure substance use with both self-report and multiple toxicological measurements. Consistent with project goals, the vast majority of ABCD participants self-reported being substance naïve at baseline, except for caffeine [12]. Recent (24 hours) substance use is assessed with breathalyzer for alcohol, urinalysis for nicotine, and oral fluid for other drugs [1, 13]. To detect substance use in the past 3 months, hair samples were collected and analyzed [14].

Hair testing measures specific drug analytes [e.g., cannabidiol, or CBD; Δ-9-tetrahydrocannabinol, or THC; 15]. Hair collection is (1) non-invasive (unlike blood), (2) not limited to recent consumption (unlike oral fluid or blood), (3) less susceptible to adulteration or dilution (unlike urine), (4) and suggests intensity of substance use [16, 17]. On average, drug metabolites are incorporated into the hair shaft after about 8 days since last use [18, 19]. Damaged or chemically-treated hair may have lower analyte concentrations, and must be considered for proper interpretation [20]. Environmental exposure from smoke, dust, or transfer from hands can also increase analyte concentrations, increasing the risk of false positives, though proper hair wash procedures can mitigate this risk [20, 21]. Mass spectrometric screening and/or confirmation of drug analytes provides high sensitivity and specificity for hair test results [22, 23].

One ABCD Study challenge is balancing study needs with participant burden and financial constraints. Toxicological assessment ranges from multiple dollars to over $200 per sample, making it cost prohibitive to test all 11,878 ABCD participants’ samples with all forms of measurement annually. Early in the ABCD study, a small percentage of participants were randomly assigned to testing with oral fluid, breathalyzer, or nicotine urine tests. Although hair was collected from 70% of participants and stored for future analysis, samples from 696 participants were selected for hair analysis based on an expert-devised evidence-based algorithm prioritizing participants according to known risk factors for substance use onset. In addition, participants who self-reported substance intake received toxicological testing during their session(s) and, as financially permissible and warranted based on the risk algorithm, by hair analysis.

We present initial ABCD cohort toxicological outcomes from findings in annual release 3.0 of the ABCD Study, consisting of baseline (9-10-year-olds), one-year (10-11-year-olds), and two-year (11-12-year-olds) data. First, we present basic descriptive toxicological and demographic information from a subsample at risk for early substance use. In this subsample, we investigated discrepancies between self-report and toxicological hair findings. Given data suggesting that adolescents underreport substance use, we hypothesized that hair analysis would identify additional substance exposure compared to self-report alone. Finally, we expected that higher scores on the risk-for-use algorithm would correspond with positive hair toxicology results.

## Material and Methods

### Participants

The ABCD Study is a 21-site longitudinal study of 11,878 participants funded by the National Institutes of Health and partner institutes. Participants were recruited predominately through school-based recruitment guided by epidemiological data [24]. The ABCD Study NDA 3.0 data release [Adolescent Brain Cognitive Development Study 25] includes toxicology results for 696 participants, and self- and parent substance use reports at the baseline, 1-year follow-up, and 2-year follow-up time points (i.e., ages ranging from 9 to 13 years).

### Measures

#### Demographics

Participants and their parents reported demographic characteristics, including the child’s sex at birth, household annual income, highest parental education, parent marital status, and race/ethnicity [26]. Some variables are social constructs requiring careful contextualization [27], limiting interpretability of these factors in this small subsample.

#### Acute On-Site Multi-Matrix Toxicological Testing

Toxicological assessment was performed on approximately 10% of randomly selected youth as well as those who reported any past-year substance use. This testing included: (1) the Dräger Drug Test® 5000 (DT5000; Dräger Inc., Houston, TX) oral fluid test consisting of a 7-panel screen for cocaine, opiates, cannabis, benzodiazepines, amphetamine, methamphetamine, and methadone; (2) a breathalyzer test for ethanol; and (3) NicAlert strips (JANT Pharmacal, Encino, CA) for urinary cotinine; to assess for past 12-72 hour substance exposure.

#### Youth Substance Use Interview

Youth participants completed a research assistant (RA)-administered substance use interview [1]. Youth were first reminded of confidentiality and asked if they had “heard of” a list of substances. If a participant had not heard of a substance, they were not asked about its use; otherwise, participants were asked about use of each major drug category, including low level use such as alcohol sipping or nicotine/cannabis puffs/tastes. Participants endorsing past-year substance use completed a detailed 12-month Timeline Followback interview about alcohol, nicotine (cigarettes, electronic nicotine delivery systems, smokeless tobacco, cigars, hookah, pipe, and nicotine replacement products), cannabis (smoked/vaped flower, smoked blunts, edibles, smoked/vaped concentrates, oral tinctures, and cannabis-infused alcohol drinks, synthetic cannabinoids), cocaine, cathinones, methamphetamine, ecstasy/MDMA, ketamine, gamma-hydroxy-butyrate, heroin, hallucinogens, psilocybin, salvia, anabolic steroids, inhalants, prescription stimulants, sedatives, and opioid pain relievers, and over the counter (OTC) cough/cold medicine use. Full reporting of substance use from ABCD’s cohort at baseline can be found in Lisdahl et al. [12].

#### Peer and Familial Substance Use Measures

Youth reported the number of their peers who used cannabis [28]. The Family History Assessment Module Screener (FHAM-S; [29]) assessed drug or alcohol use problems of any biological family member of the youth, as reported by the parent/guardian. Parents/guardians completed the Adult Self Report [30], including 3 items on parent substance use (drinking too much, daily cigarette use, and illicit substance use).

#### Childhood Behavior Checklist (CBCL)

The CBCL [30] contains questions about youths’ behavioral and mental health; it is completed by the parent/guardian. Normed externalizing symptoms (e.g., behavioral or social disturbances) were calculated.

### Procedures

Participants were assessed by trained RAs at each of the ABCD 21 research sites. Parents provided written informed consent, while youth assented. Youth participated with one parent/guardian at baseline and year 2 follow-up for approximately seven hours of behavioral, neuroimaging, and biological assessment over one or two sessions. At year 1 follow-up, the visit was typically 3 hours. Parents and youth took part separately in the protocol, with all aspects approved by a centralized Institutional Review Board.

Hair samples were collected by RAs according to standardized procedures and stored for each participant who agreed to collection and had any head hair longer than 1 cm. RAs noted hair length and color, and hair damage or dyeing, and shipped hair to Psychemedics (Culver City, CA). After receipt, hair was trimmed to 3.9 cm from the root, providing an average window of substance use detection of 3-months. Hair was enzymatically digested by a patented procedure {Hill V., 2013 #1012} and screened byFDA-cleared immunoassays or by Laboratory Developed Tests via LC-MS/MS (for Δ-9-tetrahydrocannabinol [THC], Δ9-tetrahydrocannabivarin [THCV], cannabidiol [CBD], cannabinol [CBN], and ethyl glucuronide [EtG]). Samples then underwent a 15-minute wash procedure with 2 mL isopropanol per 12 mg hair, followed by three 30-min phosphate buffer washes and, for most drugs, an additional two 60-min phosphate buffer washes [21]. This reduced possible false-positive test results and exposure via external contact only, as hair washing removes most environmental drug residue. Each presumptive positive sample was confirmed and quantified by LC-MS/MS or GC-MS/MS analysis [21, 31]. Samples were tested to the limit of detection (e.g., for THCCOOH, 0.02 pg/mg) to maximize sensitivity (see Table 1) due to the low expected degree of exposure in this young cohort.

**Table 1.**
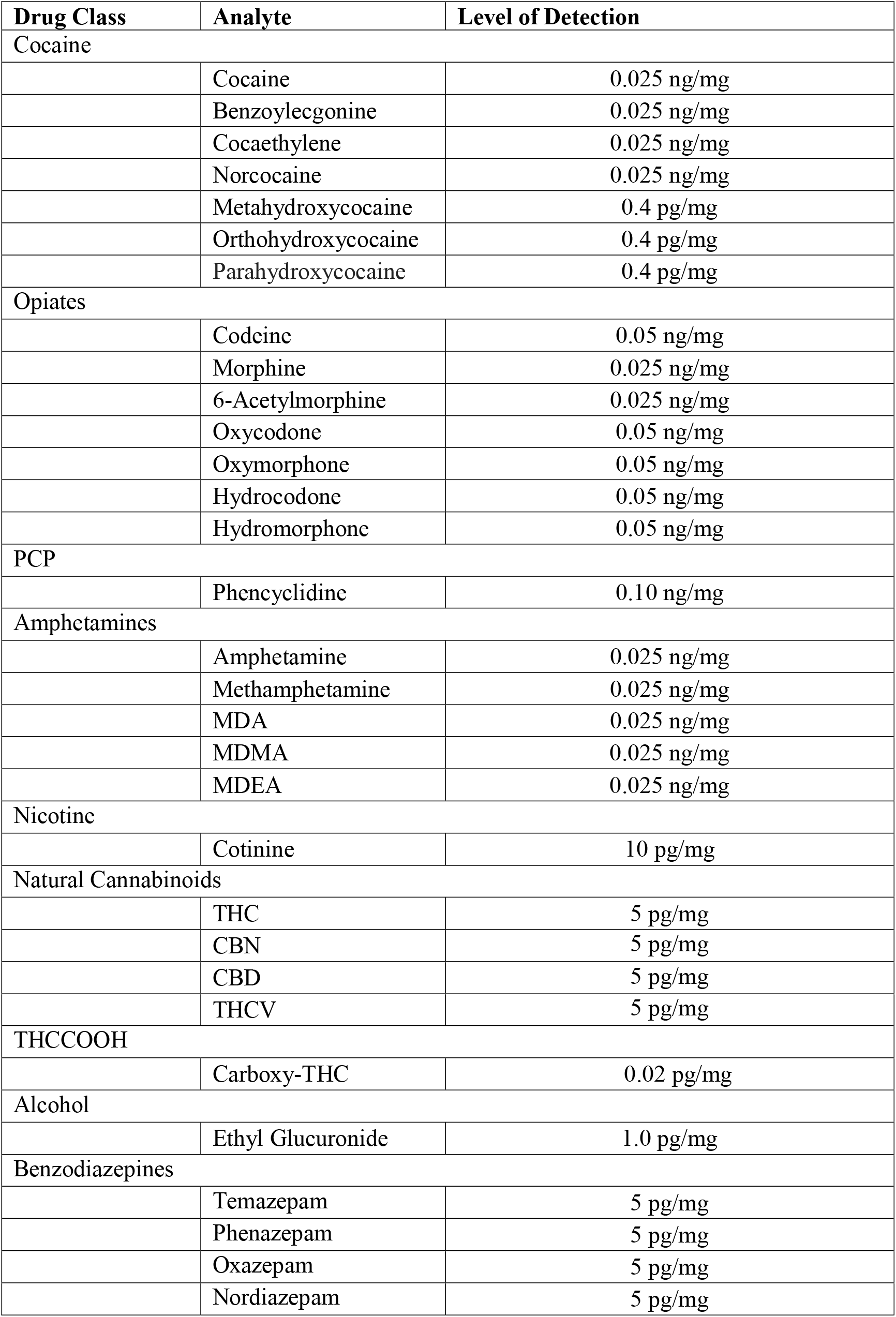
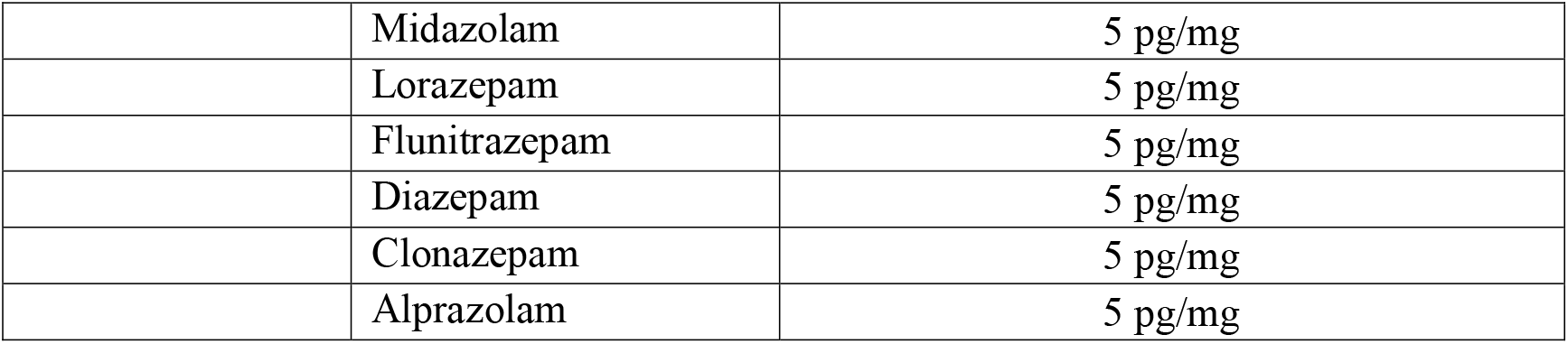
Limits of detection for each drug analyte.

An adequate hair sample was 100 mg (typically 3.9 cm in length) to complete all screening and confirmatory analyses. For some samples with insufficient weight/mass, testing was performed for as many analytes as possible, with untested screening or confirmation analyte results labeled “Quantity Not Sufficient” (QNS). Tested drug classes included cocaine, opioids, phencyclidine, amphetamines, cannabinoids, alcohol, nicotine, fentanyl, and benzodiazepines. Other analytes were available as follow-up for some samples, hence fewer results were reported for these analytes (e.g., meta-hydroxycocaine).

#### Hair Test Risk Algorithm

An evidence-based algorithm prioritized high-risk participants for hair testing (details in Supplement 1, full algorithm at https://osf.io/mtp4k/). The algorithm includes any lifetime substance use of cannabis, tobacco, prescription medications for non-medical purposes, cannabidiol (CBD), any positive oral fluid or breathalyzer test, youth reporting they will try cannabis soon, youth reporting curiosity about trying cannabis, peer cannabis use, externalizing symptoms from the CBCL, parental self-report of drug use or drinking too much alcohol, parental self-reported amount of tobacco used per day, any biological family member with drug use problems, any biological family member with alcohol use problems, youth reported drinking without parents’ approval, age and prior positive hair tests. Certain variables (i.e., positive acute toxicology test; reported CBD use) automatically selected the hair samples for testing, while having a prior positive hair test was scored at the maximum value to ensure sample analysis. Further, participants who reported more than experimental drug use (e.g., more than a puff of nicotine; more than a sip of alcohol) were selected for hair analysis.

### Statistical Analysis

SPSS 26 was utilized for all statistical analyses. Demographics (mean, SD, range, or percentage) were examined for the whole sample and by each positive substance. Both initial hair screening results as well as hair confirmation results are reported. Demographic and self-reported substance use group differences between those with positive confirmed hair toxicology results and those with negative results were assessed with chi-square and t-tests. While these analyses include assessment for differences in race/ethnicity, we note that race/ethnicity itself is a proxy for a number of different variables (e.g., educational opportunities; socioeconomic status; acculturation [32]); thus, race/ethnicity findings are reported but not discussed. Finally, we used Spearman’s rank-order correlation to evaluate the association between the evidence-based risk algorithm and positive toxicology results.

## Results

### Descriptive Data

#### Demographics

Hair analysis was performed on 696 samples, including two samples for each of 19 participants. Mean±SD participant age was 10.65±1.02 years (9-13.3), with 47.1% (n=328) from females. Full demographic details are presented in Table 2, with 61.4% Baseline samples, 16.9% Year 1 Follow-Up, and 21.8% Year 2 Follow-Up. No participants reported using substances recreationally at a level that should produce positive hair tests.

**Table 2.** Demographics of ABCD participants with hair toxicology tests, and those with positive and negative hair test results indicating substance exposure.

|  | **Hair**<br>**Toxicology**<br>**Subsample**<br>**(n=696)** | **Confirmed**<br>**Positive via**<br>**Hair**<br>**(n=116)** | **Confirmed**<br>**Negative via**<br>**Hair**<br>**(n=580)** | **Significant**<br>**Difference**<br>**by**<br>**Confirmed**<br>**Result** |
| --- | --- | --- | --- | --- |
| Age (years; range: 9-13) | 10.65 years<br>(SD = 1.02) | 10.8<br>(SD = 1.00) | 10.62<br>(SD = 1.02) | -- |
| Female | 47.1% | 37.1% | 49.1% | $p = .02$ |
| Parents Married | 63.1% | 45.7% | 66.6% | $p < .001$ |
| *Race/Ethnicity:* | | | | $p < .001$ |
| Asian | 0.7% | 0% | 0.8% |  |
| Black | 5.3% | 12.9% | 3.8% |  |
| Hispanic | 17.8% | 17.2% | 17.9% |  |
| White | 64.4% | 54.3% | 66.4% |  |
| Other/Multiple | 11.8% | 16.4% | 11.0% |  |
| *Parent Education:* | | | | $p < .001$ |
| Less than High School Diploma | 3.1% | 6.9% | 2.4% |  |
| High School/GED | 7.6% | 13.8% | 6.4% |  |
| Some College | 29.0% | 43.1% | 26.2% |  |
| Bachelor's Degree | 27.3% | 12.1% | 30.3% |  |
| Post Graduate Degree | 32.9% | 24.1% | 34.7% |  |
| *Yearly Household Income:* | | | | $p < .001$ |
| <\$50,000 | 26.0% | 44.8% | 22.2% | |
| ≥\$50,000 and <\$100,000 | 27.4% | 20.7% | 28.8% | |
| ≥\$100,000 | 40.2% | 27.6% | 42.7% | |
| *Time Point:* |  |  |  | -- |
| Baseline | 61.4% | 56.0% | 62.2% |  |
| Year 1 Follow-Up | 16.9% | 21.5% | 16.0% |  |
| Year 2 Follow-Up | 21.8% | 22.4% | 21.7% |  |

Notes: If a significant difference ( $p < 0.05$ ) was found for those with positive and negative hair tests, the p value is listed; sum totals of categories which are less than 100% are due to participants reporting “Don’t Know” or “Refuse to Answer”.

#### Hair Toxicology Results

Positive screening results were obtained for 131 of 696 hair samples, with ten samples not confirming and five samples having an insufficient amount of hair for confirmation, yielding 116 hair samples that were confirmed positive. Of these, 97 were positive for one, 14 for two, two for three drug classes and two for four drug classes. Frequency of positive results by drug class is reported in Table 3.

**Table 3.** Frequency of Screening and Confirmation Positives by Drug Class.

|  | **Positive**<br>**Screening**<br>**Results**<br>**(N=128)** | **% of Sample** | **Positive**<br>**Confirmation**<br>**Results**<br>**(N=116)** | **% of Sample** |
| --- | --- | --- | --- | --- |
| **Cocaine** | 13 | 1.9% | 13 | 1.9% |
| **Opiates** | 2 | 0.3% | 2 | 0.3% |
| **PCP** | 0 | 0% | -- | -- |
| **Amphetamines** | 64 | 9.2% | 59 | 8.5% |
| **THCCOOH** | 25 | 3.6% | 25 | 3.6% |
| **Natural**<br>**cannabinoids** | 25 | 3.6% | 22 | 3% |
| **Cannabis**<br>**(either)^** | 41 | 5.9% | 37 | 5.3% |
| **Ethyl**<br>**glucuronide** | 11 | 1.6% | 9 | 1.3% |
| **Cotinine** | 28 | 4% | 19 | 2.7% |
| **Benzodiazepines** | 0 | 0% | -- | -- |
| **Fentanyl** | 0 | 0% | -- | -- |

Notes: Frequency of each drug class is reported; % of Sample refers to the total number of hair samples available (n=696); # QNS refers to the number of samples which screened positive but did not have sufficient quantity for confirmation analysis and so were assumed positive (and included in the Positive Confirmation column); 53 participants whose hair was positive for amphetamines also reported having a prescription for amphetamines; 11-nor-9-carboxy-tetrahydrocannabinol (THCCOOH) is a $\Delta^9$ -tetrahydrocannabinol (THC) metabolite; Natural cannabinoids refers to phytocannabinoids (THC, cannabidiol (CBD), cannabinol (CBN), or $\Delta^9$ -tetrahydrocannabivarin (THCV)); no drug class is mutually exclusive; PCP phencyclidine, ETG ethyl glucuronide.
^Cannabinoids (either) indicates a hair sample that is positive for either THCCOOH or Natural Cannabinoids or both

#### Hair Toxicology Relative to Self-Reported Prescription/OTC Medication Use

We evaluated the data for reported prescription or OTC medications that may explain positive toxicology results; 64 participants with positive hair tests also self-reported recent prescription or OTC medication use (primarily amphetamine or methylphenidate). For positive amphetamines tests, 47 of 58 (81%) self-reported prescriptions, accounting for their positive amphetamine findings. An additional eight participants prescribed amphetamine or methylphenidate were *not* positive for amphetamines in hair. No other reported medications explained any positive result. Seventy-two of 696 (10.3%) of participants’ results identified substance exposure unexplained by self-reported prescription or OTC medication use.

#### Hair Toxicology Relative to Other Toxicology Results and Self-Report

All 116 participants with positive results were assessed for other toxicology and self-report data, as shown in Table 4. Full results by drug class, with specific analyte information, are available in Supplemental Table 2. Overall, 51.7% of participants with positive hair results reported some level of substance use, including experimentation through puffing nicotine or cannabis or sipping alcohol. When comparing hair results to self-report using a paired t-test, results indicated a significantly higher rate of substance use based on hair testing than self-report (t_115_ = 10.36, *p* <0.001).

**Table 4.** Acute Toxicology and Self-Report Results in Participants with Positive Hair Toxicology Results

Toxicology Results
| Method | Number Tested | Findings |
| --- | --- | --- |
| Oral Fluid | 43 | 33 for amphetamines; no other drugs |
| NicAlert | 6 | 1 indication of recent nicotine use |
| Breathalyzer | 19 | No indications of alcohol use |
| Self-Report | 116 |  |
| Alcohol sip |  | 54 |
| Full alcohol drink |  | 1 |
| Nicotine puff |  | 14 |
| More than nicotine puff |  | 5 |
| Cannabis puff |  | 4 |
| More than cannabis puff |  | 2 |
| Inhalants |  | 1 |
| Non-prescription amphetamine |  | 1 |

#### Multiple Time Points of Hair Toxicology Results

Of the 19 participants whose hair was tested on more than one year of the study, five (26.3%) had at least one positive hair test and one (5.3%) was positive for the same drug both years. Fifteen participants (78.9%) had consistent results years 1 and 2, whether positive or negative, and four (21.1%) had inconsistent results, three were positive for amphetamines or cannabis one year, and the fourth positive for cannabis one year and cannabis and nicotine the other year. Two of these four were negative at Baseline and positive at Year 2 Follow-Up, while the other two were positive at Baseline only.

#### Group Differences between Positive and Negative Hair Samples

Participants with positive hair tests were more likely to be male (*χ*^2^ = 5.65, df = 1, *p* = 0.02, d = 0.18), have less educated parents (*χ*^2^ = 37.71, df = 4, *p* < 0.001, *v* = 0.11), lower income (*χ*^2^ = 26.93, df = 2, *p* < 0.001, *v* = 0.14), be black relative to white (*χ*^2^ = 20.05, df = 4, *p* < 0.001, *v* = 0.08), and have unmarried parents (*χ*^2^ = 18.49, df = 1, *p* < 0.001, d = 0.33). Those with positive hair tests (POS) did not differ by age or assessment time point from those with negative hair tests (NEG). In assessing alcohol sipping behavior, NEG participants were significantly more likely than POS participants to report sipping alcohol (*χ*^2^ = 16.63, df = 1, *p* < 0.001, d = 0.31), even after controlling for demographic differences as listed above (β = -.149, t_649_ = -2.92, p = 0.004). In contrast, POS and NEG group members did not differ in self-reported low-level nicotine (cigarette or ENDS) (*χ*^2^ = 2.54, df = 1, *p* = 0.11, d = 0.12) or cannabis use (*χ*^2^ = 1.46, df = 1, *p* = 0.23, d = 0.09). In POS participants, 11.2% (n = 13) reported using any drug as more than a sip or puff, compared to 10.2% (n = 59) of NEG participants (*p* = 0.74).

#### Hair Test Risk Algorithm

Spearman’s rank-order correlation assessed the relationship between a confirmed positive hair test and the hair test risk algorithm score. There was a significant positive correlation between hair risk score and positive toxicology results (*r*_*s*_ = 0.23, *p* < 0.001, d = 0.47).

## Discussion

Self-report of substance use is limited by intentional and unintentional misreporting. This may be even more the case in youth engaged in a healthy development study, though accuracy of report is understudied in adolescence and, particularly, pre-adolescence. This novel investigation assessed substance use self-report and objective hair toxicology analysis that permits a larger 3-month substance use history window. Primarily, our findings show that 10.3% of youth in this sample are exposed to substances in higher amounts and/or greater frequency than they are reporting. Given use of a wash procedure to greatly reduce environmental exposure and mass spectrometric confirmation of all drug-positive results, it is likely that exposure is through personal substance use. Youth could also be timing their use to be outside the detection window for acute toxicology assessments. Second, the evidence-based risk algorithm likely predicted youth who are more likely to have positive hair toxicology results. On balance, given this was a carefully curated algorithm and thus a non-random sample, the general prevalence of actual substance use among 9-13 year-olds is still uncertain, indicating greater toxicology testing of hair is needed both within the ABCD sample and in substance use research more broadly.

Results suggest that some youths are likely using substances that are detectable on highly specific hair analyses but denying it when queried. Even in youth who report some low-level substance use (e.g., alcohol sipping or nicotine puffing), based on their hair results, some participants appear to minimize self-reported use. Several dozen participants reported substance use levels as more than a sip or puff at baseline (ages 9-10), suggesting not all youth deny substance use on interview [12]. It is important to note that hair toxicology may not detect low level substance use [14], though detection of even single use occasions is possible [33] and measurement was assessed at the limit of detection to maximize sensitivity. Also, some youth may unknowingly consume a substance (e.g., believe they are vaping flavoring rather than nicotine, or smoke cannabis laced with a novel psychoactive substance) and thus unintentionally misreport their use. Toxicology tests and specifically hair tests with their much longer window of drug detection, provide the best means of capturing substance use onset. While results here suggest that the current hair selection algorithm for targeted testing of high-risk youth is useful, this does not guarantee that other ABCD youth have not also started using substances; nor are results representative of the full, generally healthy sample, as participants were not randomly selected for testing. Future years of ABCD data collection plan to broaden acute drug screens to all participants and analyze a larger subset of hair samples from participants, including participants randomly selected without regard to their hair risk score. Such expanded testing will better detail the effects of the onset of substance use and the general rate of substance use in a healthy development study.

Interestingly, several risk factors for early substance use onset were more commonly identified in youth whose hair tested positive: being male [34] and of lower socioeconomic status [35]. Greater rates of male substance use is also consistent with the broader ABCD cohort, where males report higher levels of early substance experimentation (e.g., sipping, puffing) than females [12]. Further, the evidence-based algorithm developed to identify early onset based on common risk factors was significantly associated with positive hair results. This may be encouraging, as it further indicates that the field accurately identified risk factors for substance initiation that may be useful for preventive efforts. Interestingly, another common risk factor, reporting early alcohol sipping, was *not* related to positive hair toxicology results. It is unclear whether this is due to early experimenters, but not frequent users, having less concerns regarding privacy, or due to the fairly common nature of sipping [12, 36], or some other factor. Together this suggests further refinement of the most salient risk factors in pre-adolescent youth are needed.

As the ABCD study is still in its early stages and youth are just entering the time of predicted early substance use onset, it will be important to carefully follow these selected youth, as well as the rest of the cohort, over time. For instance, the present data contained hair analysis results from multiple years from the same participants in only 3.3% of the hair analysis pool. Of those, only two youth were positive for at least one substance both years. As more hair is collected over time, those who were positive for a substance will be tested again, allowing for longitudinal hair analyses.

Hair samples are beneficial for confirming substance exposure over long periods of time. However, hair cannot reveal date of last use, nor does it suggest the exact product used (e.g., cannabis flower v. dabs), route of administration (e.g., vaping v. smoking), frequency of use, co-or simultaneous-use, or other patterns (e.g., weekday v. weekend use). Importantly, use of hair samples does not negate the need for collection of other metrics, including self-report. Environmental factors (e.g., parental smoke) should also be considered as potential means of contaminating hair samples, though use of hair washing protocols should significantly lessen this risk [20, 21]. Such factors are likely key to understanding the full impact of substance initiation on cognitive and other outcomes, and so it is important to collect self-report and objective substance measurement whenever possible. Hair samples also were correlated with self-reported use, urine, and oral fluid tests in another study [37].

### Study Limitations

Together, findings from the present analyses suggest robust, objective measurement of substance use is needed to ensure accuracy of self-report of substance initiation. However, these results are not without limitations. First, hair toxicology analyses may not be sensitive to low levels of substance use despite the low limits of detection in this study (Table 1). In addition, hair samples may not identify early experimentation of use, as a minimal detectable dosage is not established for some drugs of abuse [33]. Though an extensive hair wash procedure was employed [21], there is the possibility of environmental drug exposure. Hair samples are also limited to the length of hair analyzed. Thus, as all hair sampled in the present study was trimmed to 3.9 cm, results reflect only the past three months of substance use. Certain youth may be more likely to decline or be unable to contribute hair samples (e.g., those with short hair “fades”, braids, or dreadlocks). In addition, there were a number of samples that did not have sufficient quantity of hair (QNS) for analysis, including five who screened positive but whose results were unable to be confirmed, suggesting we could underestimate the total number of positive results. Finally, given the financial cost of hair analyses, samples selected were restricted to those most likely to be underreporting substance use. For this reason, findings are not generalizable to the general population, or even to the overall ABCD cohort. More broad-based, randomized hair sampling is needed to better understand prevalence of underreporting in healthy developing youth.

In summary, initial hair toxicology results from the ABCD cohort suggest that 9-13 year-olds identified as most likely to initiate substance use may underreport substance use. An alarming 10.3% of high-risk youth with assayed samples were positive for at least one substance, with some participants using three or more drug classes, including nicotine, cannabis, and cocaine, although few reported even minimal use. Thus, to accurately determine the consequences of substance use in youth, greater use of robust hair samples is needed.

## Data Availability

All data is available through NIMH data archive.

## Acknowledgements

Data used in the preparation of this article were obtained from the Adolescent Brain Cognitive Development (ABCD) Study (https://abcdstudy.org), held in the NIMH Data Archive (NDA). This is a multisite, longitudinal study designed to recruit more than 10,000 children aged 9-10 and follow them over 10 years into early adulthood. The ABCD Study is supported by the National Institutes of Health and additional federal partners under award numbers U01DA041048, U01DA050989, U01DA051016, U01DA041022, U01DA051018, U01DA051037, U01DA050987, U01DA041174, U01DA041106, U01DA041117, U01DA041028, U01DA041134, U01DA050988, U01DA051039, U01DA041156, U01DA041025, U01DA041120, U01DA051038, U01DA041148, U01DA041093, U01DA041089, U24DA041123, U24DA041147. A full list of supporters is available at https://abcdstudy.org/federal-partners.html. A listing of participating sites and a complete listing of the study investigators can be found at https://abcdstudy.org/consortium_members/. ABCD consortium investigators designed and implemented the study and/or provided data but did not necessarily participate in analysis or writing of this report. This manuscript reflects the views of the authors and may not reflect the opinions or views of the NIH or ABCD consortium investigators. The ABCD data repository grows and changes over time. The ABCD data used in this report came from ABCD Release 3.0 (DOI: 10.15154/1519007). This work was also supported by K08 DA050779 (PI: Wade) and T32 AA013525 (PI: Riley/Tapert to Wade).

