## Supplement 1 for "Substance Use Onset in High-Risk 9-13 Year-Olds in the ABCD Study"

**Hair Test Risk Algorithm**

As noted in the primary document, a significant challenge for toxicology testing in the ABCD Study is the cost to test each participant. As such, acute toxicology testing (i.e., urine, breath, saliva) was reserved for those who reported use and randomization procedures for an additional percentage of youth (see Lisdahl et al., 2018).

Regarding hair sampling in particular, hair was collected from all youth who were willing to allow sample collection and whose hair was long enough to sample (>1cm in length). All samples are securely stored in temperature-controlled rooms and locked cabinets at the data collection site. Samples were collected from approximately 70% of participants each year; however, the cost of analyzing the samples is high and was not possible for all samples. Thus, members of the ABCD Substance Use Work Group (SFT, KML, FH) and an outside consultant toxicology expert (MAH) used the existing substance use risk literature to devise an algorithm to prioritize analysis of samples for those most likely to have substance intake. This approach relied on prior studies (e.g., (Gorka et al., 2014; Heron et al., 2013; Maggs et al., 2015)) and reviews (e.g., Clark & Winters, 2002; Donovan & Molina, 2011) of risk factors in youth who transition to substance use. Weights were assigned based on study aims (e.g., identifying cannabis use onset in particular and its sequelae). Variables collected in the ABCD Study populated the algorithm (see  Supplemental Table 1).

Biannually, the Data Analytics, Informatics, and Resource Center (DAIRC) of the ABCD Study calculates the hair test risk score for each participant with hair samples available. Samples from those with higher scores on the algorithm, more recent hair collections, participants who previously tested positive, and participants who tested positive on acute toxicological assessment are prioritized. Data collection sites are requested to ship the selected samples to the Psychemedics laboratory for analysis. Psychemedics analyzes the samples as described in the full paper, with screen, confirmation, and quantified analyte results provided back to the DAIRC for upload to the NDA data release.

Supplemental Table 1. Sample prioritization algorithm variables and scoring.

| **Variable** | **Source Questionnaire** | **Points Conferred** |
|---|---|---|
| Lifetime Substance Use | Timeline Follow-Back | Up to 15 pts (1 pt per endorsed variable) |
| Specific lifetime use of cannabis | Timeline Follow-Back | 10 pts |
| Positive saliva toxicology | Draeger DrugTest 5000 | 10 pts |
| Intention to use cannabis soon | PATH Questionnaire | Up to 5 pts |
| Curiosity about using cannabis | PATH Questionnaire | Up to 4 pts |
| How many peers use cannabis | Peer Substance Use | Up to 4 pts |
| Elevated externalizing t-score | Achenbach's Childhood Behavior Checklist | 3 pts |
| Parental drug use | Achenbach's Adult Self Report | Up to 2 pts |
| Parental drinking too much | Achenbach's Adult Self Report | Up to 2 pts |
| Parental tobacco use | Achenbach's Adult Self Report | 1 pt |
| Any biological relative with history of Alcohol Use Disorder | Family History | 1 pt |
| Any biological relative with history of drug use disorder | Family History | 1 pt |
| Drinks alcohol without parent's approval | Achenbach's Childhood Behavior Checklist | Up to 2 pts |

| | | |
|---|---|---|
| Smokes, chews, or sniffs tobacco | Achenbach's Childhood Behavior Checklist | 2 pts |
| Uses drugs recreationally | Achenbach's Childhood Behavior Checklist | 2 pts |
| Medical use of CBD | Medication Form | 5 pts |
| Positive breathalyzer screen | Breathalyzer Toxicology Report | 5 pts |
| Prior positive | Psychemedics Hair Sample Report | Automatically identified for sending |
| | | Maximum: 76 pts |

Notes: Details on specific question items and measures can be reviewed in Lisdahl et al., 2018. Variables with multiple possible options have different scores for each corresponding response, with the highest score reflecting the highest risk response; these items are denoted as being scored as "up to ___ points".


Clark, D. B., & Winters, K. C. (2002). Measuring risks and outcomes in substance use disorders prevention research. *J Consult Clin Psychol*, *70*(6), 1207-1223. https://doi.org/10.1037//0022-006x.70.6.1207

Donovan, J. E., & Molina, B. S. G. (2011). Childhood Risk Factors for Early-Onset Drinking. *Journal of Studies on Alcohol and Drugs*, *72*(5), 741-751. https://doi.org/10.15288/jsad.2011.72.741

Gorka, S. M., Shankman, S. A., Olino, T. M., Seeley, J. R., Kosty, D. B., & Lewinsohn, P. M. (2014). Anxiety disorders and risk for alcohol use disorders: the moderating effect of parental support. *Drug Alcohol Depend*, *140*, 191-197. https://doi.org/10.1016/j.drugalcdep.2014.04.021

Heron, J., Maughan, B., Dick, D. M., Kendler, K. S., Lewis, G., Macleod, J., Munafo, M., & Hickman, M. (2013). Conduct problem trajectories and alcohol use and misuse in mid to late adolescence. *Drug Alcohol Depend*, *133*(1), 100-107. https://doi.org/10.1016/j.drugalcdep.2013.05.025

Lisdahl, K. M., Sher, K. J., Conway, K. P., Gonzalez, R., Feldstein Ewing, S. W., Nixon, S. J., Tapert, S., Bartsch, H., Goldstein, R. Z., & Heitzeg, M. (2018). Adolescent brain cognitive development (ABCD) study: Overview of substance use assessment methods. *Dev Cogn Neurosci*, *32*, 80-96. https://doi.org/10.1016/j.dcn.2018.02.007

Maggs, J. L., Staff, J., Patrick, M. E., Wray-Lake, L., & Schulenberg, J. E. (2015). Alcohol use at the cusp of adolescence: a prospective national birth cohort study of prevalence and risk factors. *J Adolesc Health*, *56*(6), 639-645. https://doi.org/10.1016/j.jadohealth.2015.02.010
