## Supplement 2 for "Substance Use Onset in High-Risk 9-13 Year-Olds in the ABCD Study"

All ABCD participants completed a substance use interview, including a 12-month Timeline Follow-Back if they reported any substance use. In addition, all participants were asked to give a sample of their hair for analysis. Ten percent of participants were randomly selected to undergo some or all of the following acute toxicology tests: breathalyzer test, urinary cotinine screen, and/ororal fluid testing. Finally, participants who reported any past year substance use were given all three acute toxicology assessments. Data reported in Table S2 show self-reported substance use, detailed hair toxicology results, and acute toxicology results in participants who had positive hair results.

**Table S2.** Positive acute toxicology results and self-reported use endorsements for participants with confirmed positive hair results, by drug class.

| **Positive result per self-report or biospecimen:** | **Cocaine (n=13)** | **Opiates  (n=2)** | **Amphetamines  (n=59)** | **THCCOOH  (n=25)** | **Natural Cannabinoids (n=22)** | **Alcohol  (n=9)** | **Nicotine  (n=19)** |
| --- | --- | --- | --- | --- | --- | --- | --- |
| Self-Reported use: |  |  |  |  |  |  |  |
| Alcohol sips | 38.5% | 50% | 40.7% | 48.0% | 59.1% | 66.7% | 52.6% |
| Alcohol full drink | 0% | 0% | 0 | 4.0% | 4.5% | 0% | 0% |
| Nicotine puffs | 15.4% | 50% | 5.1% | 16.0% | 22.7% | 11.1% | 21.1% |
| Nicotine more than puff | 7.7% | 0% | 0% | 4.0% | 4.5% | 11.1% | 5.3% |
| Cannabis puff | 7.7% | 50% | 0% | 12.0% | 9.1% | 11.1% | 10.5% |
| Cannabis more than puff | 7.7% | 50% | 0% | 4.0% | 4.5% | 11.1% | 5.3% |
| Other drugs | 0% | 0% | 0% | 0% | 0% | 0% | 0% |
| Hair analyte confirmation ^b^: |  |  |  |  |  |  |  |
| Cocaine ^d^ | 13/13 |  |  |  |  |  |  |
| Benzoylecgonine | 12/12 |  |  |  |  |  |  |
| Norcocaine | 2/11 |  |  |  |  |  |  |
| Oxymorphone ^c^ |  | 2/2 |  |  |  |  |  |
| Amphetamines ^e^ |  |  | 52/59 |  |  |  |  |
| Methamphetamine |  |  | 9/51 |  |  |  |  |
| THCCOOH |  |  |  | 25/25 | 10/11 |  |  |
| THC |  |  |  | 9/10 | 21/22 |  |  |
| CBN |  |  |  | 3/5 | 5/7 |  |  |
| CBD |  |  |  | 5/5 | 8/11 |  |  |
| THCV |  |  |  | 0/2 | 0/4 |  |  |
| ETG |  |  |  |  |  | 9/9 |  |
| Cotinine |  |  |  |  |  |  | 18/19 |
| 3-hydroxycotinine |  |  |  |  |  |  | 6/11 |
| Norcotinine |  |  |  |  |  |  | 1/1 |
| Nornicotine |  |  |  |  |  |  | 8/8 |
| Draeger oral fluid test ^f^: |  |  |  |  |  |  |  |
| Amphetamine | 1/1 ^a^ | 0/0 | 30/30 ^a^ | 0/3 | 2/3 ^a^ | 0/0 | 2/4 ^a^ |
| NicAlert ^g^ | 0/4 | 0/0 | 0/2 | 1/3 | 1/3 | 0/0 | 1/1 |

Notes: 11-nor-9-carboxy-tetrahydrocannabinol = THCCOOH, ∆9-tetrahydrocannabinol = THC, cannabidiol = CBD, cannabinol = CBN, ∆9-tetrahydrocannabivarin = THCV; ETG=ethyl glucuronide

^a^ Positive results are consistent with the youth's prescription medication

^b^ Variation in analyte n due to insufficient quantity of hair (e.g., norcocaine; THCV)

^c^ Other opioids tested for (fentanyl, codeine, morphine, 6-acetylmorphine, hydrocodone, hydromorphone, and oxycodone) were negative for all.

^d^ Other cocaine metabolites tested for (cocaethylene, metahydroxycocaine, orthohydroxycocaine, and parahydroxycocaine) were all negative.

^e^ Other amphetamines tested for (MDA, MDEA, and MDMA) were all negative.

^f^ Seven drug classes (cocaine, opiates, cannabis, benzodiazepines, amphetamine, methamphetamine, and methadone) were tested as described in the primary manuscript. Only drug classes with a positive are displayed here.

^g^NicAlert is a urinary cotinine screen. Positive results on NicAlert here indicae urinary cotinine results at a level which indicates recent, personal nicotine exposure.
